# Single-cell RNA sequencing identifies subtypes of cancer-associated fibroblasts in early and late stages of mycosis fungoides

**DOI:** 10.1101/2025.09.07.25335167

**Authors:** Courtney M. Johnson, Weishan Li, Soroosh Solhjoo, Vrinda Madan, Iman Ali, Kalvin Nash, Stephanie Hicks, Winston Timp

**Author notes:** **Corresponding author:** Courtney M. Johnson, Johns Hopkins University, Department of Dermatology, 1550 Orleans Street, CRBII, Suite 207, Baltimore, MD 21287.

## Abstract

Mycosis fungoides (MF), the most common cutaneous T-cell lymphoma, is characterized by infiltration of malignant T-cells into the skin. While early-stage disease (IA-IIA) follows an indolent course, approximately 25% of patients progress to late-stage disease (IIB-IVB) with significantly worse prognosis and a median survival of 1-5 years. Identifying which early-stage MF patients are at high risk for disease progression remains difficult. This may be affected by the complex interaction between malignant tumor cells and the tumor microenvironment. Increased numbers of cancer-associated fibroblasts (CAF) are found in early-stage MF and have been shown to support tumor growth, but the subtypes have not yet been classified in lesional MF tissue. We performed single-cell RNA sequencing on skin samples from healthy individuals, early-stage MF patients, and late-stage MF patients to investigate the fibroblast population. Analysis of the highly differentially expressed genes in the fibroblast populations revealed nine distinct subclusters, comprising five major types: ECM/structural, vascular/metabolic, immune-modulatory, antigen-presenting, and developmental fibroblasts. Stage-specific differences revealed that the vascular/metabolic subcluster was enriched in early-stage MF, while the ECM/structural subcluster was enriched in late-stage MF. An antigen-presenting subcluster, a novel and underrecognized subtype, was identified by its high expression of MHC class II genes and pathways essential for antigen processing and presentation of exogenous peptides. These inappropriate antigen-presenting fibroblasts may play a role in chronic T-cell exhaustion seen in late-stage diseases. Further studies with additional patient samples will validate these findings and clarify the role of these subtypes in diagnosing and predicting the outcome of mycosis fungoides.

**Translational Relevance:** Our findings reveal heterogeneity among fibroblasts in mycosis fungoides (MF) skin lesions, highlighting potential prognostic biomarkers and therapeutic targets with direct implications for improving the risk assessment and treatment of advanced disease. Using single-cell RNA sequencing, nine transcriptionally distinct fibroblast subclusters were identified that are involved in ECM remodeling, immune modulation, and metabolic adaptation. Examining both early- and late-stage MF disease, in a racially diverse patient cohort, offers new insights into how fibroblast composition changes during disease progression and across demographics. Additionally, we characterize an antigen-presenting fibroblast population in MF that may play an underappreciated role in the chronically exhausted T cells of advanced-stage disease.

## Introduction

Mycosis fungoides (MF), the most common form of cutaneous T-cell lymphoma, is a clonal disorder of skin-homing memory T cells, with an incidence of 6.4 cases per million individuals _[1]_. Early-stage MF (IA-IIA) has an indolent disease course, with a 10-year overall survival rate of 52-88% and mortality rates comparable to those of age-matched controls _[2]_. Over time, about 25% of patients progress to late-stage disease (IIB-IVB), during which malignant T cells infiltrate the lymph nodes, blood, and visceral organs, resulting in a median survival of 1-5 years _[2, 3]_. Identifying which patients will progress to late-stage disease remains a challenging and unpredictable task.

Understanding the role of the supporting stroma, which includes fibroblasts and other immune cells, can provide critical insights into the prognosis of MF. Cells in the tumor microenvironment (TME) play a vital role in tumor progression by offering structural support, suppressing immune responses, modulating therapeutic resistance, and influencing tumor evolution _[4, 5]_. Fibroblasts are increasingly recognized as active regulators of tumor biology [6]. In addition to maintaining the extracellular matrix (ECM), fibroblasts play pivotal roles in modulating stromal-epithelial interactions during tumor invasion. They promote tumorigenesis by driving T helper 2 (Th2) responses, which dampen anti-tumor immunity, while simultaneously suppressing T helper 1 (Th1) responses, which are critical for immune surveillance [7–9]. Cancer-associated fibroblasts (CAFs) are activated fibroblasts that differ in phenotype, function, and location from normal fibroblasts _[9]_. CAFs have been shown to support tumor growth in various solid and hematological malignancies, with specific subtypes of CAFs associated with poor clinical outcomes _[10, 11]_.

Single-cell RNA sequencing (scRNA-seq) studies have identified CAF subtypes in various malignancies, highlighting their roles in immune suppression, extracellular matrix (ECM) remodeling, and metabolic adaptation _[12]_. Yet, the single-cell resolution of CAFs in MF remains poorly defined. Furthermore, much of our knowledge about CAFs in MF relies on bulk transcriptomic analyses and fibroblast isolation techniques that do not distinguish between different fibroblast subpopulations _[13–16]_. Moreover, the significances of disease stage and racial diversity on fibroblast composition have yet to be investigated. Here, we present a pilot study employing scRNA-seq to characterize fibroblast heterogeneity in MF, identifying distinct fibroblast subclusters across various disease stages and self-reported racial backgrounds. By sampling a diverse patient cohort that includes both early- and late-stage disease, we provide a comprehensive view of the contributions of fibroblasts to MF progression, uncovering novel subtype-specific functions in ECM remodeling, immune modulation, and antigen presentation. Our findings enhance the current understanding of MF-associated fibroblasts, offering new insights into their potential roles in disease progression and therapeutic response. Such findings could provide prognostic insights and enable personalized treatment plans.

## Methods and Materials

The Johns Hopkins Institutional Review Board approved the study protocol for specimen collection and sequencing (IRB00331637). With informed consent, four MF patients and one patient without MF were recruited from the Johns Hopkins Cutaneous Lymphoma Clinic. All patients diagnosed with MF had biopsy-proven disease, as confirmed by a dermatopathologist. For scRNA-seq, patches, thick plaques, or tumors were selected, and a 4 mm punch size was used to collect tissue. For the patient without MF, normal-appearing, unaffected skin from a double-covered truncal location was selected for collection. Disease staging for patients with MF was determined by the EORTC/WHO staging guidelines, in which patients with early-stage MF were defined as stages IA to IIA, and those with late-stage MF as stages IIB or higher. Patient characteristics and clinical disease staging are outlined in **Figure 1A**.

**FIGURE 1.**
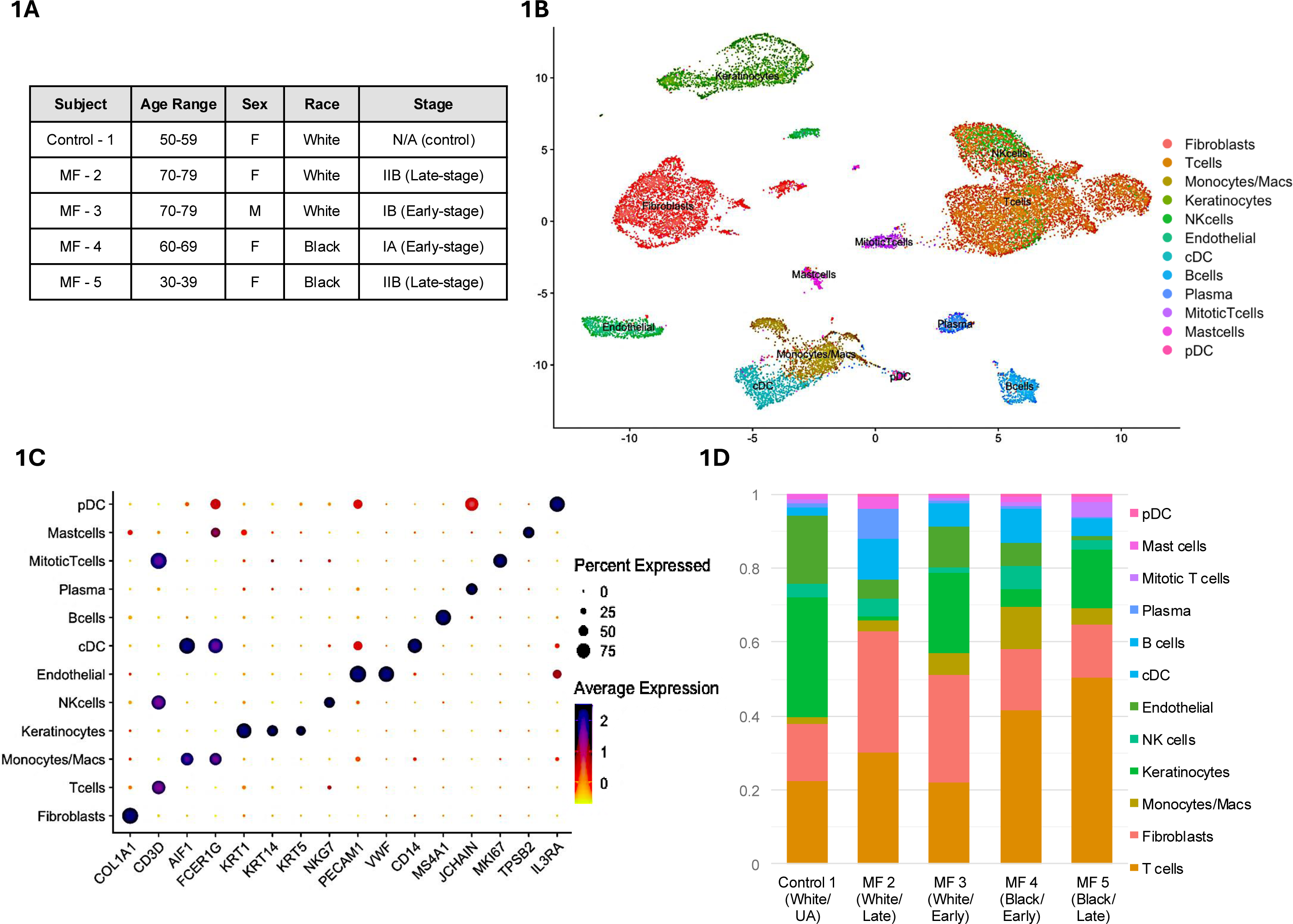
scRNA-sequencing results of skin from mycosis fungoides (MF) and healthy control (HC) samples. Transcriptomes of 28,902 cells (1,906 from HC (n=1) and 26,996 from MF (n=4) skin samples) were analyzed using the Seurat v4 pipeline. Data were log-normalized and scaled; principal component analysis (PCA) was performed on the top variable genes, and unsupervised clustering was generated using a shared nearest neighbor (SNN) modularity optimization algorithm (resolution = 0.5). Differential expression analysis used the Wilcoxon rank-sum test with Bonferroni correction (adjusted *p* < 0.05). Cluster identities were assigned based on canonical marker expression. Dot size represents the percentage of cells within a cluster expressing the indicated gene, while dot color represents the average expression level (SCTransform-normalized, log1p-transformed Pearson residuals), ranging from 0 (yellow) to 2 (dark blue). A) Clinical data of all five patients included in the study. B) Unsupervised clustering, represented by Uniform Manifold Approximation and Projection (UMAP), shows dimensional reduction of reads from all five samples combined. C) Dot plot showing the percentage and average expression of signature genes for clusters grouped by gene marker: Fibroblasts (COL1A1), T-cells (CD3D), Natural Killer Cells (NKG7), Keratinocytes (KRT1, KRT14, KRT5), Monocytes/Macrophages (AIF1, FCER1G), Endothelial Cells (PECAM1, VWF), B-cells (MS4A1), Plasma Cells (JCHAIN), Mitotic T-cells (CD3D, MK167), Mast Cells (FCER1G, TPSB2), Conventional Dendritic Cells (CD14), And Plasmacytoid Dendritic Cells (IL3RA). D) Stacked bar chart illustrating the relative proportion of identified cell populations across the control and four MF samples

Freshly biopsied skin samples underwent immediate tissue dissociation. This is followed by single-cell capture, library preparation, and single-cell RNA sequencing with 10X Genomics. A more detailed outline of the methods and sequencing analysis can be found in the supplemental data. Descriptive data from the genomic analysis are summarized in **Supplemental Table S1**. The data were then analyzed using Seurat (v4) for quality control, filtration, and data integration. To correct for batch effects and emphasize shared biological signals across varying patient samples, we applied horizontal data integration using the Harmony function, which aligns similar cell populations across different datasets without removing biologically meaningful variation _[17, 18]_. The top 3000 highly expressed genes were identified in the merged Seurat object, and a UMAP of cell clusters was created based on the harmonized principal components (PCs).

Differential gene expression was conducted on each cluster using the *FindMarkers* function (Wilcoxon rank-sum Test). The threshold result was adjusted for p-value > 0.05 and positive differentially expressed genes. Cluster-specific differentially expressed genes were kept, and canonical gene markers were used to annotate the clusters. Fibroblasts were identified by their expression of *COL1A1* and were subsequently subclustered for further analysis based on the top 3000 variable genes. Gene Ontology enrichment analysis was performed at P<0.05.

## Results

Unsupervised clustering analysis of 28,902 cells from MF and control skin samples revealed twenty-nine transcriptionally distinct cell clusters as visualized on the UMAP **Figure 1B** and **Supplemental Figures S1-S3**. Significant cell populations were identified using canonical gene markers, with each cell population exhibiting transcriptionally distinct cellular signatures, highlighting the complexity of the collected samples (**Figure 1C, Supplemental Table S2**).

Figure 1D highlights both the conserved shifts in cell composition in MF disease and patient-specific variability, with some patients exhibiting marked immune cell infiltration or fibroblast expansion, possibly reflecting disease stage or host factors. T-cells (*CD3D*), the predominant immune cells, were enriched in MF samples, with a median of 36% (range, 21-50%) compared to the control (22%), consistent with the known infiltration of T-cells in MF skin lesions. Keratinocytes (*KRT1*, *KRT14*, *KRT5*) were markedly reduced in MF skin (median 10%, range 9-22%) relative to controls (33%), suggesting epidermal dysregulation or disruption. A baseline of fibroblasts was noted across all samples, with the proportion of fibroblasts in MF samples (median 23%, range 14-32%) similar to that of the control (16%). Monocytes/ Macrophages (*AIF1*, *FCER1G*), Natural Killer Cells (*NKG7*), Endothelial Cells (*PECAM1*, *VWF*), B-cells (*MS4A1*), Plasma Cells (*JCHAIN*), Mitotic T-cells (*CD3D*, *MK167*), Mast Cells (*FCER1G*, *TPSB2*), Conventional Dendritic Cells (*CD14*), and Plasmacytoid Dendritic Cells (*IL3RA*) were variably represented across all samples. These results demonstrate the significant heterogeneity of the TME in MF lesions.

A population of 4,672 fibroblasts, identified by COL1A1 expression, was further isolated into nine transcriptionally distinct subclusters of fibroblasts based on differential gene expression (Figure 2A, **Supplemental Figure S4**). The largest subtype, subcluster 0, accounted for 46% of fibroblasts and was enriched in late-stage MF (average 72%) compared to early-stage (average 23%) and control (54%) (Figure 2B). Notably, subcluster 5 exhibited the highest expression in the control patient (average 15%) with decreasing expression in early-stage (11%) and late-stage (0.01%) patients, suggesting a functional shift across MF stages (Figure 3A-B). Fibroblast heterogeneity varied across patient samples. Patient 4 (early-stage, Black) demonstrated the highest diversity, containing all fibroblast subclusters, while Patient 2 (late-stage, White) had the lowest diversity, representing only five subclusters (Figure 2B**, Supplemental Figure S5).**

**FIGURE 2.**
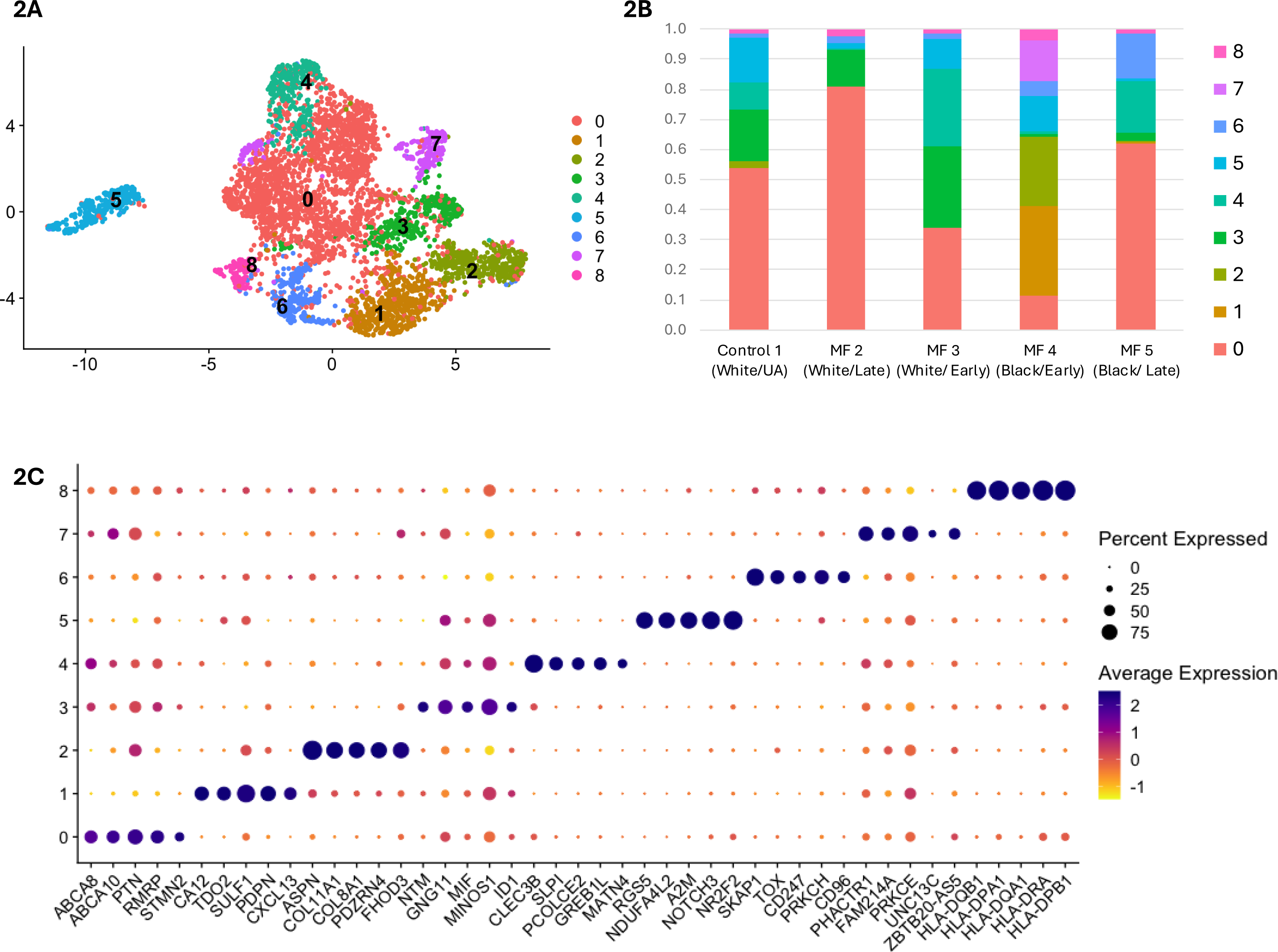
scRNA-sequencing identifies distinct fibroblast subclusters in mycosis fungoides (MF). Transcriptomes of 4,672 COL1A1+ cells (186 from HC (n=1) and 4,486 from MF (n=4)) were analyzed to resolve fibroblast heterogeneity. Dot size represents the percentage of cells in each subcluster expressing the indicated gene, while dot color indicates average expression (SCTransform-normalized values, log1p-transformed Pearson residuals, ranging from –1 [yellow] to 2 [dark blue]). A) Unsupervised clustering visualized by UMAP reveals nine transcriptionally distinct fibroblast subclusters across all samples. B) Stacked bar chart showing the relative proportion of each fibroblast subcluster by individual patient sample C) Dot plot displaying the top five differentially expressed genes (DEGs) for each subcluster, highlighting unique transcriptional profiles.

**FIGURE 3.**
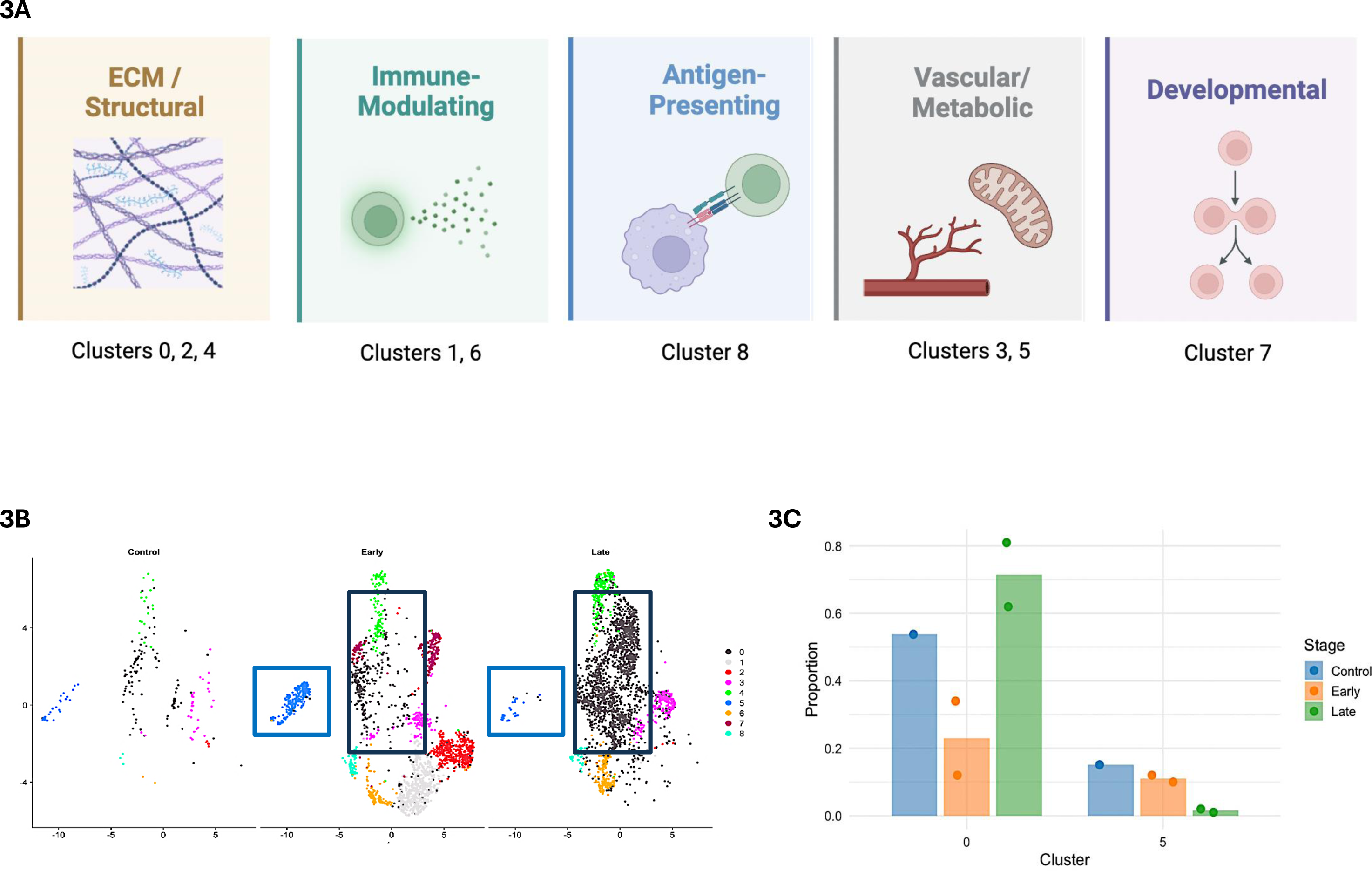
Cancer-associated fibroblast (CAF) phenotypes identified in fibroblast subclusters. Differential expression was performed using the Wilcoxon rank-sum test (log₂ fold-change ≥ 0.25, adjusted p < 0.05, Bonferroni correction). Gene ontology analysis was conducted using the top 50 differentially expressed genes per subcluster (hypergeometric test with Benjamini–Hochberg correction, q < 0.05). A) Schematic illustration summarizing five major CAF phenotypes identified among the nine fibroblast subclusters based on differentially expressed gene patterns and gene ontology enrichment results. B) Dot plot showing the distribution of the nine fibroblast subclusters across healthy control, early-stage mycosis fungoides (MF), and late-stage MF samples. The boxes highlight clusters 0 and 5. C) Bar heights represent the mean proportion of fibroblasts in clusters 0 and 5 for each disease stage (Control, MF Early, MF Late). Individual dots denote values from each of the five patient samples, illustrating inter-patient variability.

To functionally characterize the fibroblast subclusters, differential expression analysis of the top genes and Gene Ontology (GO) enrichment analysis (GO_Biological_Process_2021) were performed to reveal distinct biological roles (Figure 2C**, Supplemental Figure S6)**. A summary of the highly differentially expressed genes and their functions in each subcluster is outlined in **Table 1**. In summary, the nine distinct subclusters demonstrate roles in major cancer-associated phenotypes, including extracellular matrix remodeling (subclusters 0, 2, 4), immune modulation (subclusters 1, 6), vascular/metabolic (subclusters 3, 5), development (subcluster 7), and antigen presentation (subcluster 8) (Figure 3A). Fibroblast subclusters demonstrated stage-specific distributions. Early-stage MF was notably enriched in subclusters associated with vascular remodeling and metabolic adaptation (subcluster 5). Conversely, late-stage MF showed significant enrichment in ECM remodeling fibroblasts (subcluster 0) (Figure 3B-C).

**TABLE 1.**
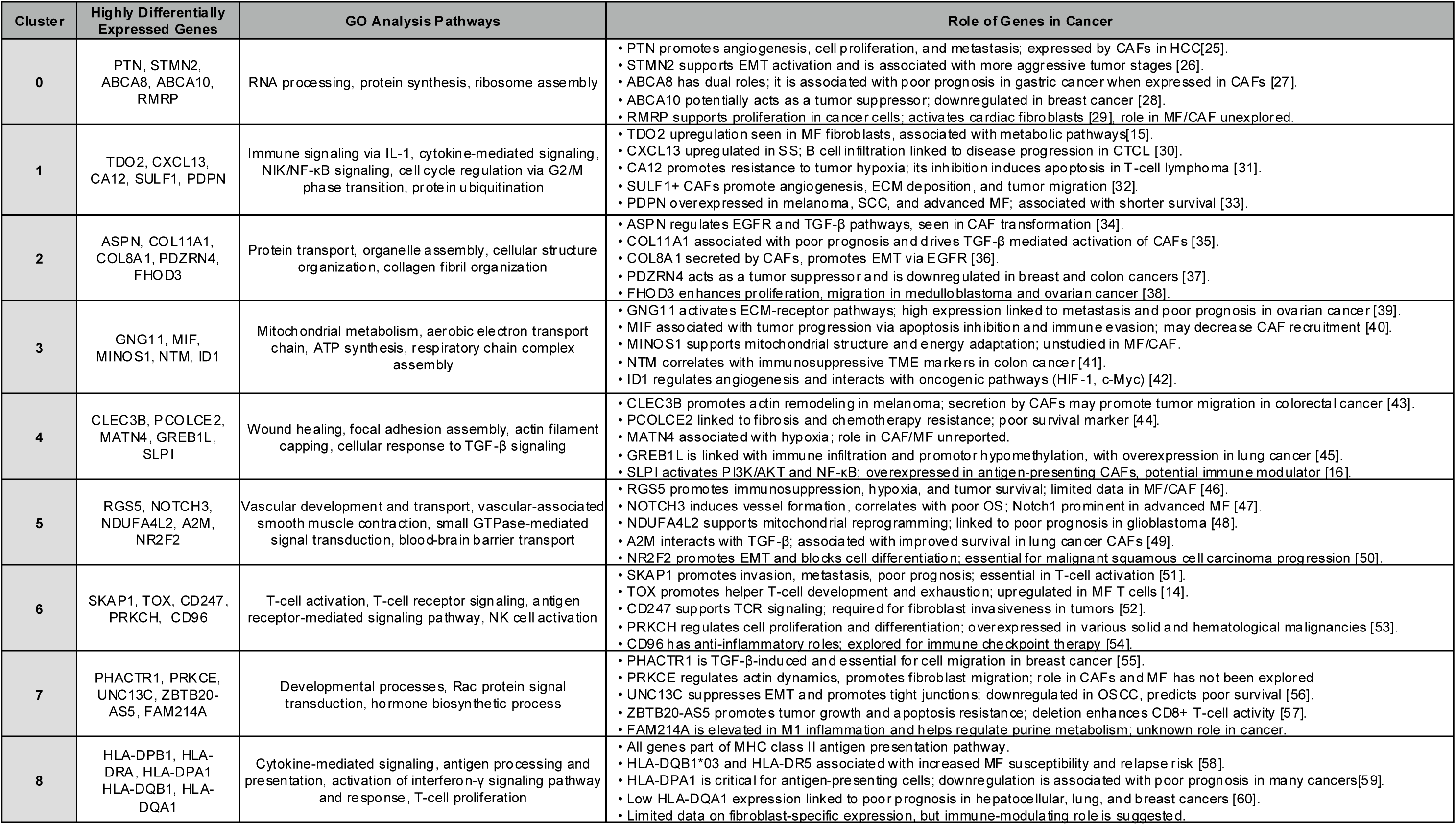
Differentially expressed genes (DEGs) and gene ontology (GO) enrichment reveal functional specialization of fibroblast subclusters. Top highly differentially expressed genes for each fibroblast subcluster were identified using the Wilcoxon rank-sum test as implemented in Seurat (log₂ fold-change ≥ 0.25, adjusted p < 0.05, Bonferroni correction). Gene ontology (GO) enrichment analysis was performed on the top 50 DEGs per cluster using a hypergeometric test with Benjamini–Hochberg correction (q < 0.05). Enriched pathways highlight distinct functional roles of fibroblast subclusters, including extracellular matrix organization, immune regulation, and cytokine signaling.

## Discussion

In this study, we employed single-cell RNA sequencing to investigate the transcriptional heterogeneity of fibroblasts in mycosis fungoides skin and identified nine distinct subclusters, each with a role in major cancer-associated phenotypes of fibroblasts. Our findings suggest that specific fibroblast populations may contribute to disease progression by shaping the tumor microenvironment and influencing immune response dysfunction. Across disease stages, we observed shifts in fibroblast subtypes, such as vascular/metabolic (subcluster 5), which were enriched in early-stage MF, whereas ECM-remodeling (subcluster 0) dominated the late stages of the disease. These shifts may reflect an evolving stromal landscape during MF progression from immune-active and metabolically dynamic early lesions to fibrotic, immunosuppressive TME in late-stage disease.

Our racially diverse patient cohort enabled an exploratory analysis of demographic differences in fibroblast composition. Notably, fibroblast subcluster 6, enriched for genes involved in T-cell signaling and immune modulation (e.g., TOX, CD247), was more prominent in the self-reported Black patient sample, where patients 4 (5%) and 4 (15%) had higher proportions compared to patients 1-3 with 1%, 2%, and 2%, respectively. In contrast, subcluster 3, which is linked to chronic inflammation and mitochondrial metabolism (e.g., MIF, MINOS1), was enriched in self-reported White patients. These findings suggest that racial differences in stromal-immune interactions may exist in MF, potentially contributing to observed disparities in disease severity, progression, or immune phenotype. Although limited by sample size, these patterns are consistent with prior reports that have shown racial variation in MF incidence, stage at diagnosis, and outcomes _[19, 20]_. Further investigation in larger, balanced cohorts using hereditary-based SNPs or other genetic markers (rather than self-reported race) is essential to clarify whether fibroblast heterogeneity contributes to these disparities and to determine whether targeting fibroblast subtypes may offer personalized therapeutic opportunities.

One of the most intriguing findings is the identification of a previously uncharacterized subset of antigen-presenting fibroblasts (subcluster 8), marked by high expression of MHC class II genes (HLA-DRA, HLA-DPB1) and enrichment for antigen-processing and interferon-γ signaling pathways. These fibroblasts may participate in non-canonical antigen presentation and modulate local immune responses by influencing the polarization of CD4+ T cells. While antigen presentation by fibroblasts can activate immune surveillance in certain contexts, within MF, persistent MHC II expression without co-stimulation may promote T-cell anergy or exhaustion, as observed in late-stage disease _[21]._ This result mirrors findings in other cancers where antigen-presenting CAFs dampen cytotoxic responses and foster tolerogenic environments _[22]_.

In addition to antigen presentation, several fibroblast subclusters expressed cytokines and chemokines that shape the immune milieu. For instance, subcluster 1 exhibited elevated CXCL13, which is implicated in the recruitment of B- and T-follicular helper cells _[23]_. In contrast, subcluster 3 expressed MIF, a cytokine known to suppress cytotoxic T-cell activity and promote chronic inflammation _[24]_. These fibroblast-derived mediators likely reinforce immune dysregulation and contribute to the spatial and functional exclusion of effective anti-tumor responses within the MF lesion.

While our study provides valuable insights into MF fibroblast biology, further work is needed to validate these subclusters functionally. Sampling led to unequal numbers of fibroblasts across samples, which we addressed by merging, normalizing, and integrating markers through computational analysis. For future studies, the use of spatial transcriptomics will enable the precise localization of these fibroblast subtypes within MF lesions, thereby elucidating their interactions with malignant T cells and other immune components. Furthermore, a larger patient cohort would help refine the stage- and demographic-associated differences in fibroblasts, facilitating further stratification of fibroblast populations based on clinical and molecular characteristics.

## Conclusion

This study provides a comprehensive single-cell transcriptomic analysis of fibroblasts in mycosis fungoides, revealing nine distinct subpopulations with diverse roles in extracellular matrix remodeling, immune modulation, and metabolic adaptation. By sampling patients across early and late disease stages and including a diverse cohort, we uncover stage- and race-associated shifts in fibroblast composition that may inform our understanding of disease biology and progression. Most notably, we identify a novel population of antigen-presenting fibroblasts that may contribute to immune dysfunction through aberrant MHC class II expression and interferon-γ–driven signaling. Together with the discovery of subclusters enriched in cytokines and immunoregulatory factors (e.g., CXCL13, MIF, TOX), our findings suggest that cancer-associated fibroblasts actively shape the immunopathology of MF beyond structural support. These insights highlight the therapeutic potential of targeting specific fibroblast subtypes or their signaling pathways to modulate the tumor microenvironment, improve immune surveillance, and ultimately alter disease trajectory in mycosis fungoides.

## Data Availability Statement

The data generated in this study are available within the article and its supplementary data files. The expression profile data analyzed in this study are pending submission to the Gene Expression Omnibus (GEO).

## ETHICS STATEMENT

All patients and control participants gave written, informed consent. The Johns Hopkins University Institutional Review Board approved this study.

## CONFLICT OF INTEREST STATEMENT

The authors state no conflict of interest.

## ACKNOWLEDGMENTS

We recognize and appreciate the funding support of the NIH K99/R00, Maryland Dermatological Society, Skin of Color Society, Dermatology Foundation, and the NIH T32 program. We want to thank Dr. Ronald Sweren for providing access to his patient panel for inclusion in this study. Finally, we would like to thank the patients who have graciously participated in this project.

## Author Contributions

**Courtney Johnson:** conceptualization (equal), data curation (lead), formal analysis (equal), investigation (lead), visualization (lead), writing – original draft (lead), funding acquisition (lead). **Weishan Li:** formal analysis (equal), visualization (supporting), validation (supporting), writing – review and editing (equal). **Soroosh Solhjoo**: validation (supporting), writing – review and editing (equal). **Vrinda Madan:** investigation (equal), writing – review and editing (equal). **Iman Ali:** investigation (equal). **Kalvin Nash:** investigation (equal). **Stephanie Hicks:** validation (supporting), writing – review and editing (equal). **Winston Timp:** conceptualization (equal), validation (supporting), writing – review and editing (equal).

## Abbreviations

scRNA: single-cell RNA
CAF: Cancer-associated fibroblasts
MF: mycosis fungoides

## Notes

### Competing Interest Statement

The authors have declared no competing interest.

### Funding Statement

This study was funded by the NIH/ NIAMS MOSAIC K99/R00, Maryland Dermatological Society, Skin of Color Society, Dermatology Foundation, and the NIH T32 program of Johns Hopkins University SOM Department of Dermatology.

### Author Declarations

Ethics committee/IRB of Johns Hopkins School of Medicine gave ethical approval for this work.

## References

1. Criscione VD, Weinstock MA: Incidence of cutaneous T-cell lymphoma in the United States, 1973-2002. Arch Dermatol 2007, 143(7):854–859.

2. Agar NS, Wedgeworth E, Crichton S, Mitchell TJ, Cox M, Ferreira S, Robson A, Calonje E, Stefanato CM, Wain EM, Wilkins B, Fields PA, Dean A, Webb K, Scarisbrick J, Morris S, Whittaker SJ: Survival outcomes and prognostic factors in mycosis fungoides/Sezary syndrome: validation of the revised International Society for Cutaneous Lymphomas/European Organisation for Research and Treatment of Cancer staging proposal. J Clin Oncol 2010, 28(31):4730–4739.

3. Kempf W, Mitteldorf C: Cutaneous T-cell lymphomas-An update 2021. Hematol Oncol 2021, 39 Suppl 1:46–51.

4. Gordon-Weeks A, Yuzhalin AE: Cancer Extracellular Matrix Proteins Regulate Tumour Immunity. Cancers (Basel) 2020, 12(11):3331. doi: 10.3390/cancers12113331.

5. Vinay DS, Ryan EP, Pawelec G, Talib WH, Stagg J, Elkord E, Lichtor T, Decker WK, Whelan RL, Kumara HMCS, Signori E, Honoki K, Georgakilas AG, Amin A, Helferich WG, Boosani CS, Guha G, Ciriolo MR, Chen S, Mohammed SI, Azmi AS, Keith WN, Bilsland A, Bhakta D, Halicka D, Fujii H, Aquilano K, Ashraf SS, Nowsheen S, Yang X, Choi BK, Kwon BS: Immune evasion in cancer: Mechanistic basis and therapeutic strategies. Semin Cancer Biol 2015, 35 Suppl:S185–S198.

6. Kalluri R: The biology and function of fibroblasts in cancer. Nat Rev Cancer 2016, 16(9):582–598.

7. Miyagaki T, Sugaya M, Suga H, Morimura S, Ohmatsu H, Fujita H, Asano Y, Tada Y, Kadono T, Sato S: Low herpesvirus entry mediator (HVEM) expression on dermal fibroblasts contributes to a Th2-dominant microenvironment in advanced cutaneous T-cell lymphoma. J Invest Dermatol 2012, 132(4):1280–1289.

8. Takahashi N, Sugaya M, Suga H, Oka T, Kawaguchi M, Miyagaki T, Fujita H, Sato S: Thymic Stromal Chemokine TSLP Acts through Th2 Cytokine Production to Induce Cutaneous T-cell Lymphoma. Cancer Res 2016, 76(21):6241–6252.

9. Tao L, Huang G, Song H, Chen Y, Chen L: Cancer associated fibroblasts: An essential role in the tumor microenvironment. Oncol Lett 2017, 14(3):2611–2620.

10. Arpinati L, Scherz-Shouval R: From gatekeepers to providers: regulation of immune functions by cancer-associated fibroblasts. Trends Cancer 2023, 9(5):421– 443.

11. Ding Z, Shi R, Hu W, Tian L, Sun R, Wu Y, Zhang X: Cancer-associated fibroblasts in hematologic malignancies: elucidating roles and spotlighting therapeutic targets. Front Oncol 2023, 13:1193978.

12. Xu Y, Su G, Ma D, Xiao Y, Shao Z, Jiang Y: Technological advances in cancer immunity: from immunogenomics to single-cell analysis and artificial intelligence. Signal Transduct Target Ther 2021, 6(1):312.

13. Aronovich A, Moyal L, Gorovitz B, Amitay-Laish I, Naveh HP, Forer Y, Maron L, Knaneh J, Ad-El D, Yaacobi D, Barel E, Erez N, Hodak E: Cancer-Associated Fibroblasts in Mycosis Fungoides Promote Tumor Cell Migration and Drug Resistance through CXCL12/CXCR4. J Invest Dermatol 2021, 141(3):619–627.e2.

14. Mehdi SJ, Moerman-Herzog A, Wong HK: Normal and cancer fibroblasts differentially regulate TWIST1, TOX and cytokine gene expression in cutaneous T-cell lymphoma. BMC Cancer 2021, 21(1):492–7.

15. Gaydosik AM, Stonesifer CJ, Tabib T, Lafyatis R, Geskin LJ, Fuschiotti P: The mycosis fungoides cutaneous microenvironment shapes dysfunctional cell trafficking, antitumor immunity, matrix interactions, and angiogenesis. JCI Insight 2023, 8(19):e170015. doi: 10.1172/jci.insight.170015.

16. Zhao Y, Li Y, Wang P, Zhu M, Wang J, Xie B, Tang C, Ma Y, Wang S, Jin S, Xu J, Li Z, Zhang X, Li L, Song X, Wang P: The cancer-associated fibroblasts interact with malignant T cells in mycosis fungoides and promote the disease progression. Front Immunol 2025, 15:1474564.

17. Ryu Y, Han GH, Jung E, Hwang D: Integration of Single-Cell RNA-Seq Datasets: A Review of Computational Methods. Mol Cells 2023, 46(2):106–119.

18. Korsunsky I, Millard N, Fan J, Slowikowski K, Zhang F, Wei K, Baglaenko Y, Brenner M, Loh P, Raychaudhuri S: Fast, sensitive and accurate integration of single-cell data with Harmony. Nat Methods 2019, 16(12):1289–1296.

19. Su C, Nguyen KA, Bai HX, Cao Y, Tao Y, Xiao R, Karakousis G, Zhang PJ, Zhang G: Racial disparity in mycosis fungoides: An analysis of 4495 cases from the US National Cancer Database. J Am Acad Dermatol 2017, 77(3):497–502.e2.

20. Hooper MJ, Veon FL, Enriquez GL, Nguyen M, Grimes CB, LeWitt TM, Pang Y, Case S, Choi J, Guitart J, Burns MB, Zhou XA: Retrospective analysis of sepsis in cutaneous T-cell lymphoma reveals significantly greater risk in Black patients. J Am Acad Dermatol 2023, 88(2):329–337.

