## Supplemental Figures for "Single-cell RNA sequencing identifies subtypes of cancer-associated fibroblasts in early and late stages of mycosis fungoides"

Supplemental Data

Courtney M. Johnson, M.D., Ph.D.[1], Weishan Li, B.S.[3], Soroosh Solhjoo, Ph.D. [2], Vrinda Madan, B.A.[1], Iman Ali, B.S.[1], Kalvin Nash, B.S.[1], Stephanie Hicks, Ph.D.[3,4,5,6] and Winston Timp, Ph.D.[4,5]

*[1]Department of Dermatology, Johns Hopkins University School of Medicine, Baltimore, MD, USA*
*[2]Department of Medicine, F. Edward Hébert School of Medicine, Bethesda, MD, USA*
*[3]Department of Biostatistics, Johns Hopkins Bloomberg School of Public Health, Baltimore, MD, USA*
*[4]Department of Biomedical Engineering, Johns Hopkins University School of Medicine, Baltimore, MD,*
*[5]Center for Computational Biology, Johns Hopkins University, Baltimore, MD, USA*
*[6]Malone Center for Engineering in Healthcare, Johns Hopkins University, Baltimore, MD, USA*

**Supplemental Table S1. Sequencing and quality control metrics**

| Sample: | Cells | Number of Reads | Mean Reads per Cell | Sequencing Saturation | Valid UMIs | Median UMI counts per cell | Median Genes per Cell |
|---|---|---|---|---|---|---|---|
| Control - 1 | 1,906 | 168,147,398 | 88,220 | 84.80% | 99.90% | 4,732 | 2,120 |
| MF - 2 | 5,016 | 167,622,092 | 33,425 | 74.20% | 99.90% | 3,642 | 1,498 |
| MF - 3 | 1,650 | 198,612,554 | 120,371 | 89.00% | 99.90% | 4,504 | 1,822 |
| MF - 4 | 11,135 | 195,693,228 | 17,575 | 34.10% | 99.90% | 4,043 | 1,984 |
| MF - 5 | 9,195 | 193,834,968 | 21,080 | 35.00% | 99.90% | 3,420 | 1,714 |

**Supplemental Table S2. Canonical gene markers are used to identify major cell populations.**

| Canonical Gene Markers | |
|---|---|
| Fibroblasts | COL1A1 |
| T-cells | CD3D |
| Natural Killer Cells | NKG7 |
| Keratinocytes | KRT1, KRT14, KRT5 |
| Monocytes/Macrophages | AIF1, FCER1G |
| Endothelial Cells | PECAM1, VWF |
| B-cells | MS4A1 |
| Plasma Cells | JCHAIN |
| Mitotic T-cells | CD3D, MK167 |
| Mast Cells | FCER1G, TPSB2 |
| Conventional Dendritic Cells | CD14 |
| Plasmacytoid Dendritic Cells | IL3RA |

**Supplemental Figure S1. Unsupervised clustering of all cell types from mycosis fungoides (MF) and healthy control (HC) skin.**

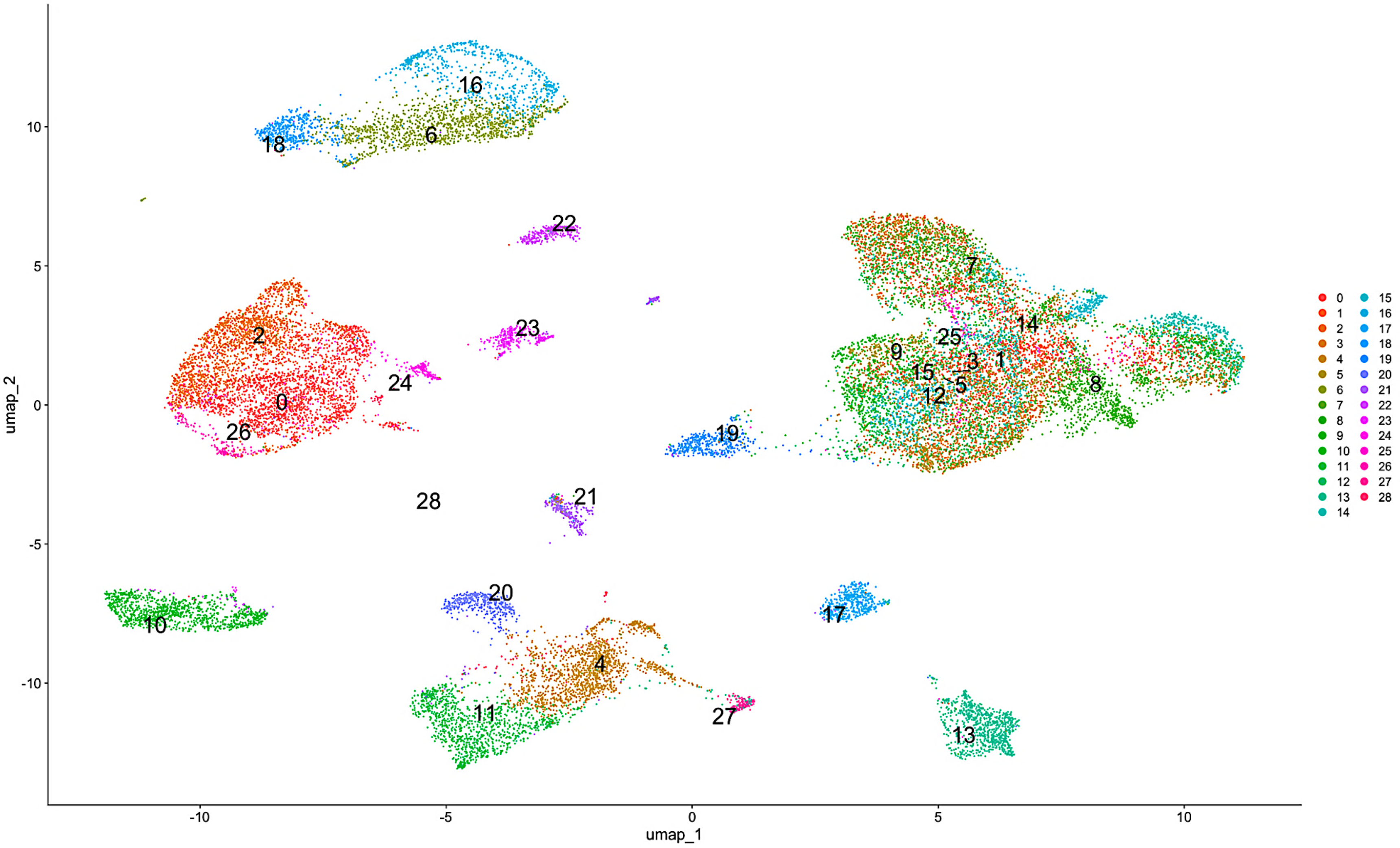

**Supplemental Figure S2. Heatmap of the top five differentially expressed genes per cluster in the combined dataset.**

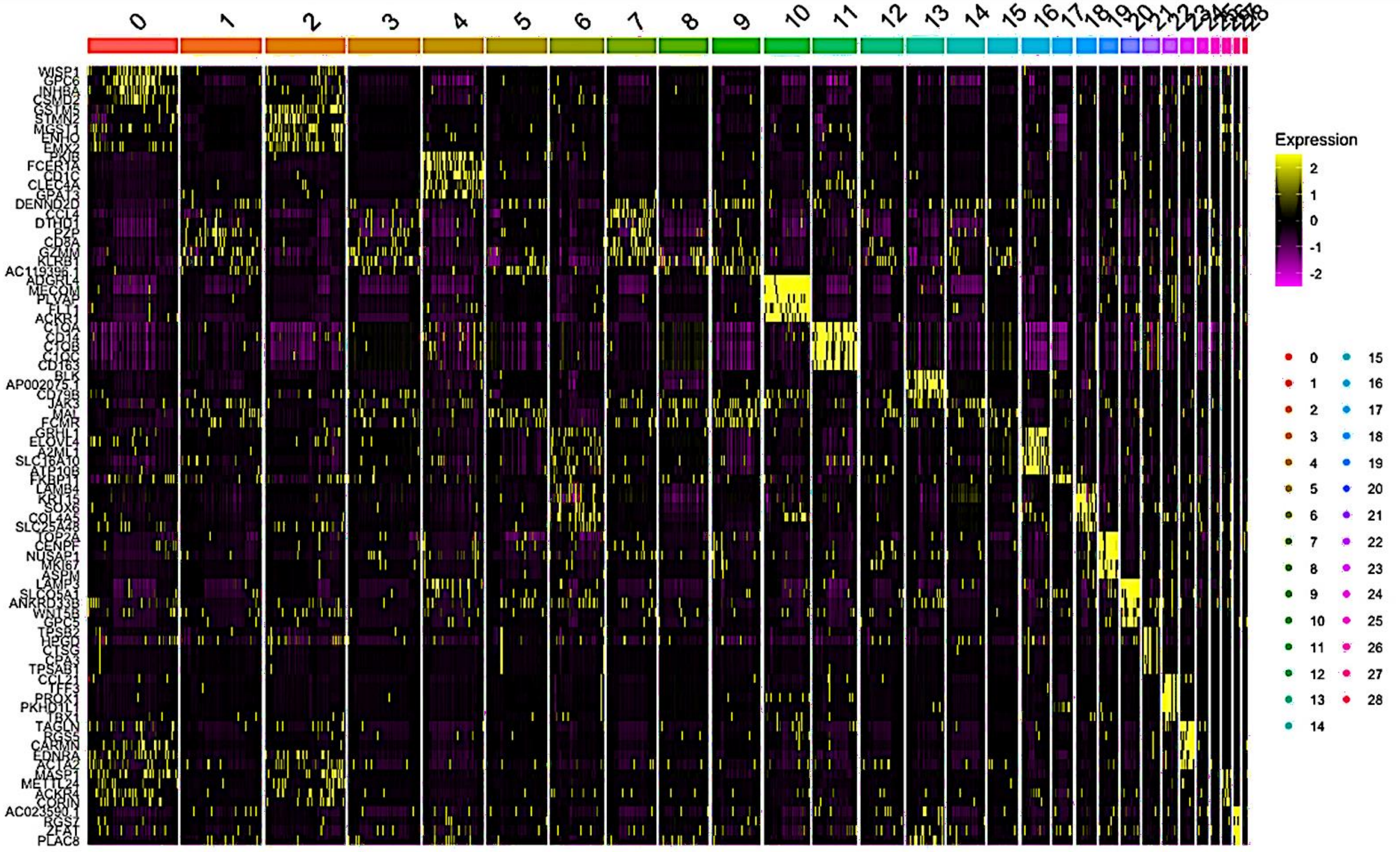

**Supplemental Figure S3. Unsupervised clustering of all cell types separated by patient sample.**

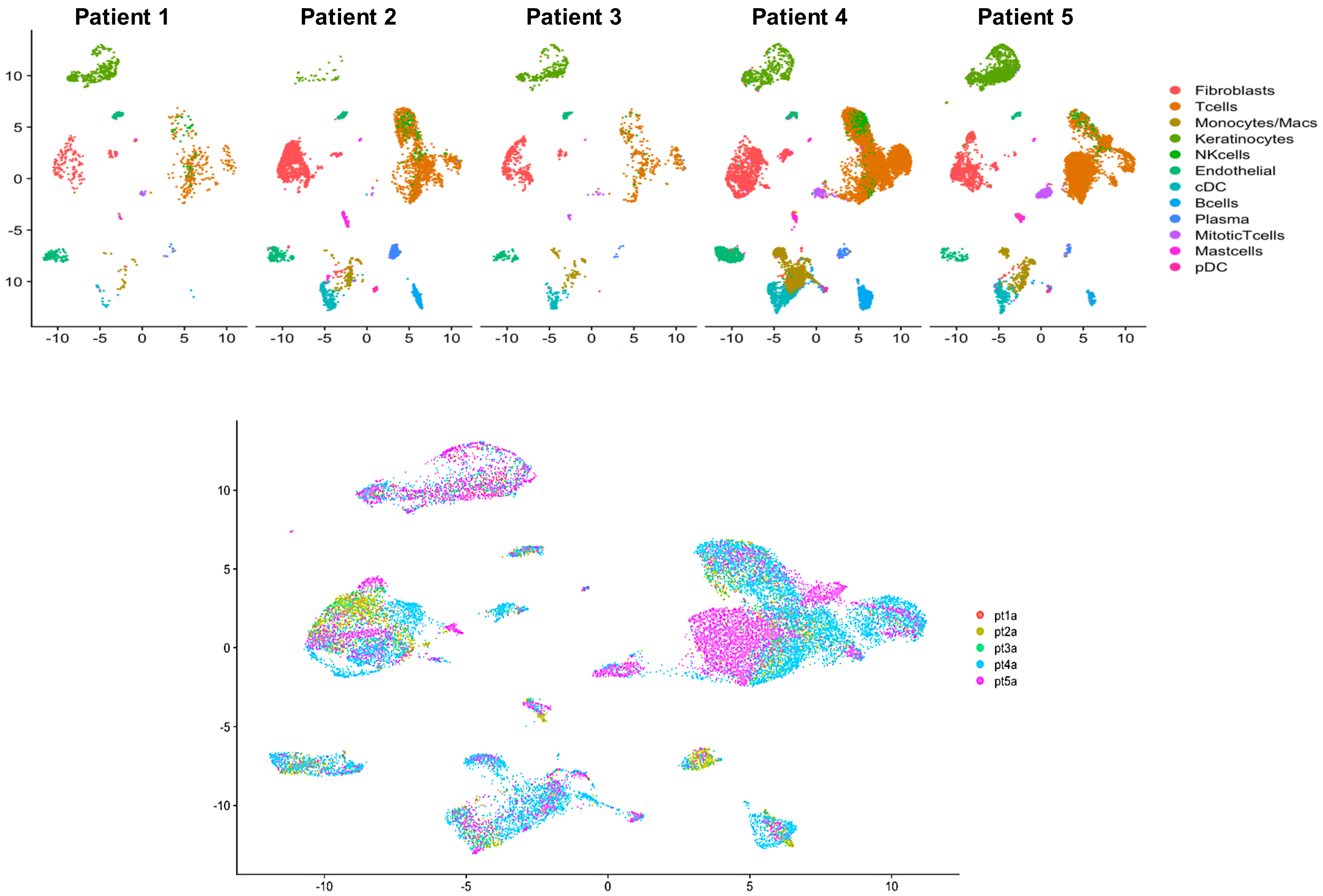

**Supplemental Figure S4. Heatmap of the top five differentially expressed genes within each fibroblast subcluster.**

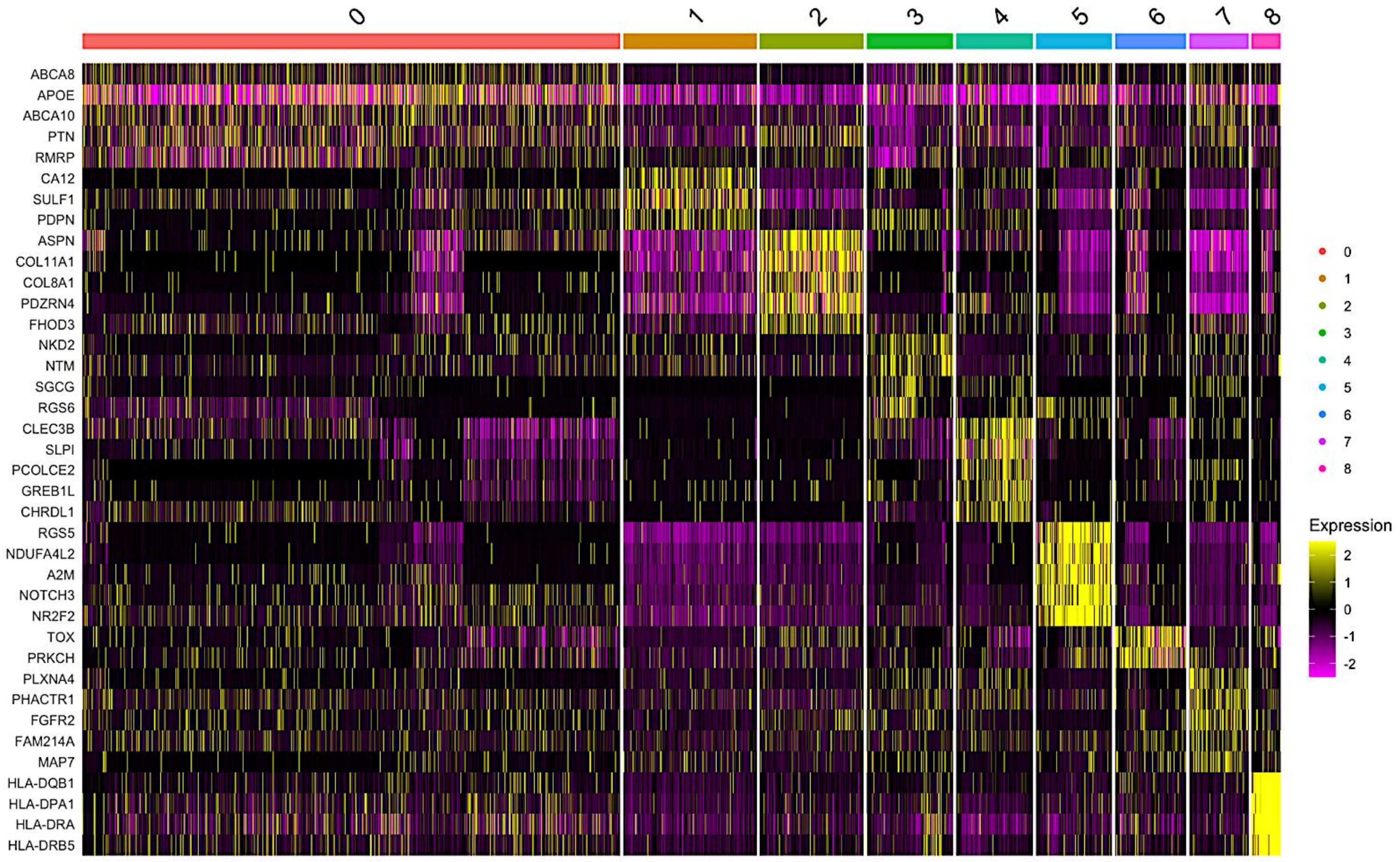

**Supplemental Figure S5. UMAP visualization of fibroblast subclusters by patient sample.**

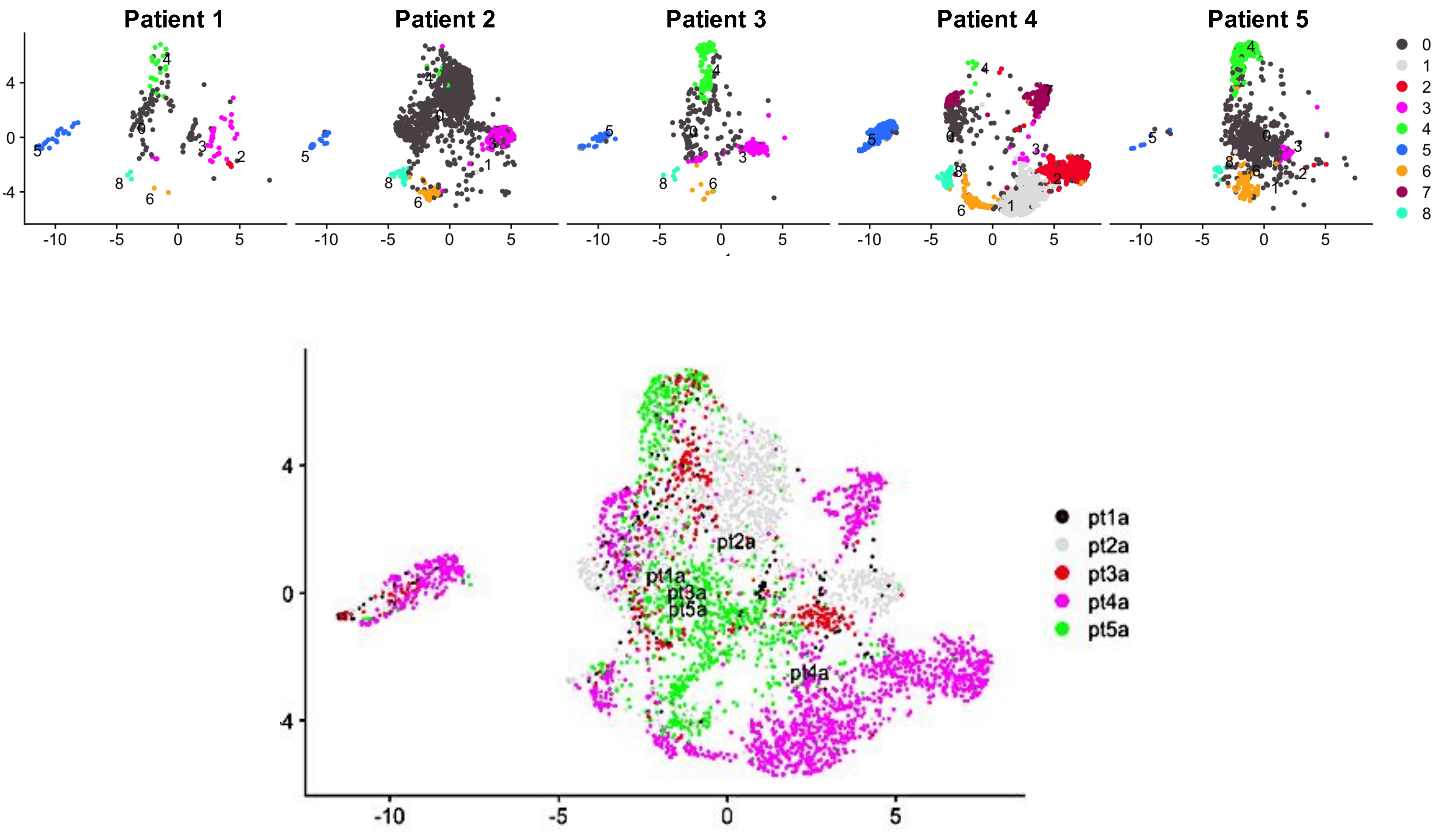

**Supplemental Figure S6. Gene ontology (GO) enrichment analysis of fibroblast subclusters**

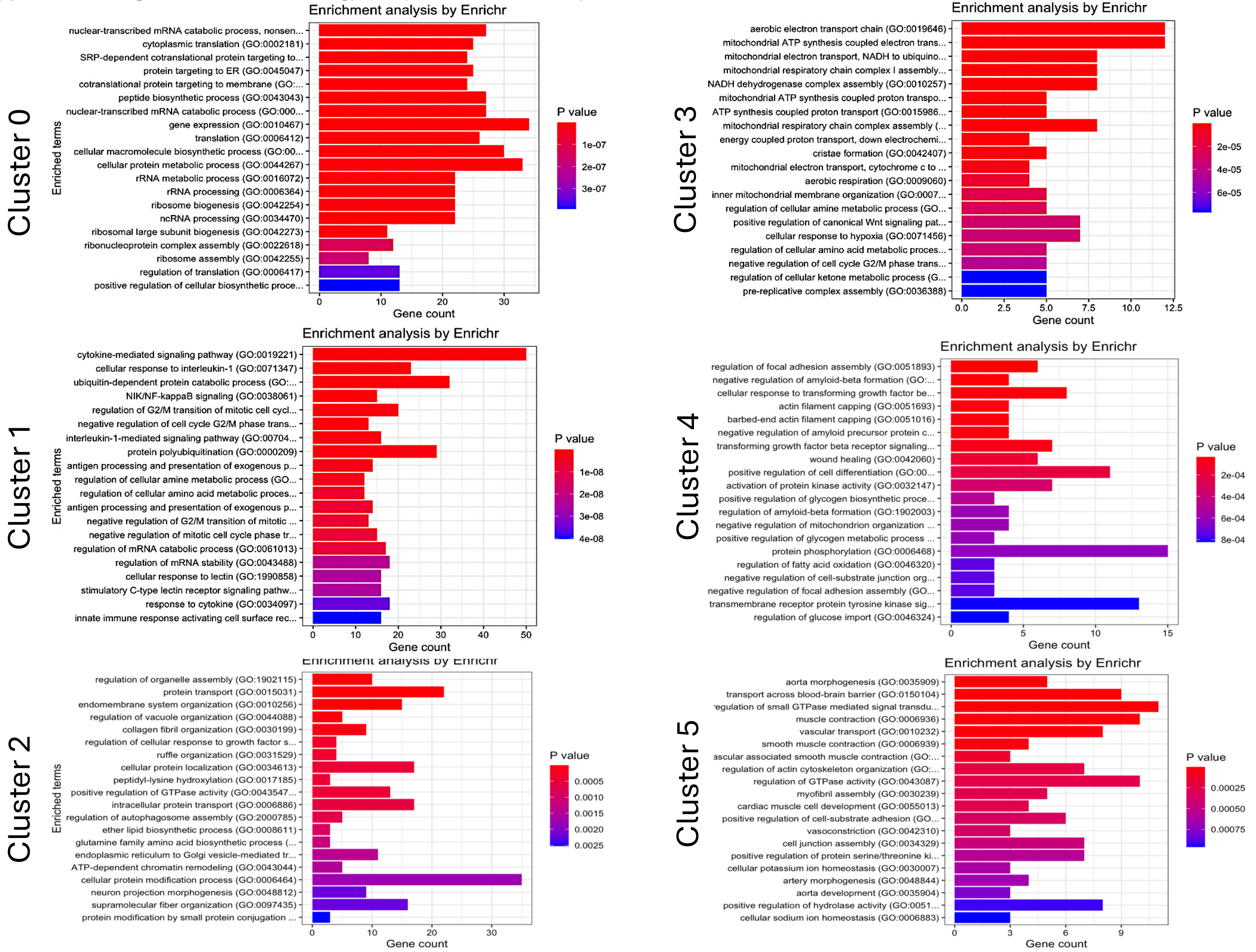

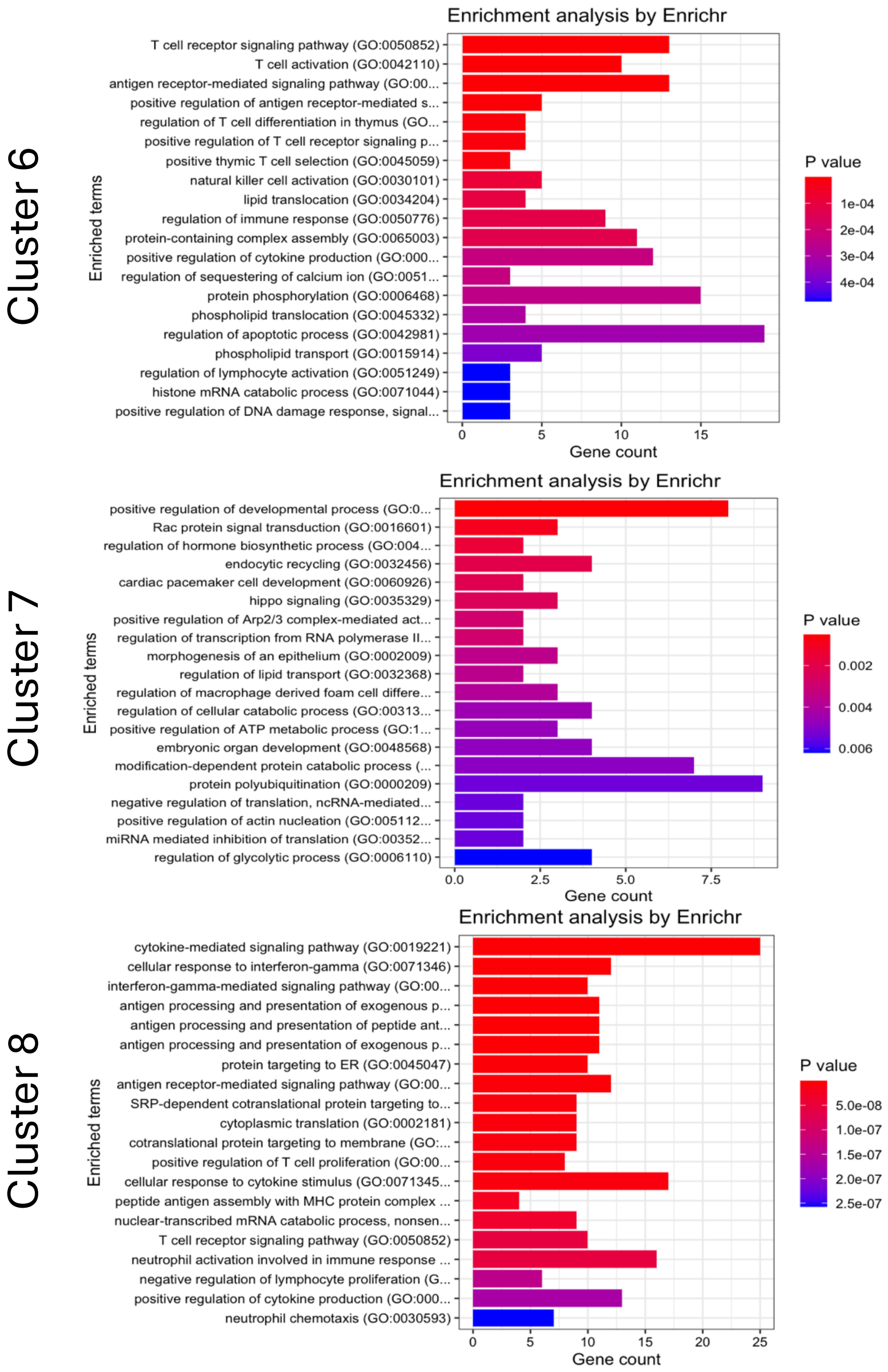
Cluster 6
Enrichment analysis by Enrichr
T cell receptor signaling pathway (GO:0050852)
T cell activation (GO:0042110)
antigen receptor-mediated signaling pathway (GO:00...
positive regulation of antigen receptor-mediated s...
regulation of T cell differentiation in thymus (GO...
positive regulation of T cell receptor signaling p...
positive thymic T cell selection (GO:0045059)
natural killer cell activation (GO:0030101)
lipid translocation (GO:0034204)
regulation of immune response (GO:0050776)
protein-containing complex assembly (GO:0065003)
positive regulation of cytokine production (GO:000...
regulation of sequestering of calcium ion (GO:0051...
protein phosphorylation (GO:0006468)
phospholipid translocation (GO:0045332)
regulation of apoptotic process (GO:0042981)
phospholipid transport (GO:0015914)
regulation of lymphocyte activation (GO:0051249)
histone mRNA catabolic process (GO:0071044)
positive regulation of DNA damage response, signal...
Enriched terms
0
5
10
15
Gene count
P value
1e-04
2e-04
3e-04
4e-04
Cluster 7
Enrichment analysis by Enrichr
positive regulation of developmental process (GO:0...
Rac protein signal transduction (GO:0016601)
regulation of hormone biosynthetic process (GO:004...
endocytic recycling (GO:0032456)
cardiac pacemaker cell development (GO:0060926)
hippo signaling (GO:0035329)
positive regulation of Arp2/3 complex-mediated act...
regulation of transcription from RNA polymerase II...
morphogenesis of an epithelium (GO:0002009)
regulation of lipid transport (GO:0032368)
regulation of macrophage derived foam cell differe...
regulation of cellular catabolic process (GO:00313...
positive regulation of ATP metabolic process (GO:1...
embryonic organ development (GO:0048568)
modification-dependent protein catabolic process (...
protein polyubiquitination (GO:0000209)
negative regulation of translation, ncRNA-mediated...
positive regulation of actin nucleation (GO:005112...
miRNA mediated inhibition of translation (GO:00352...
regulation of glycolytic process (GO:0006110)
Enriched terms
0.0
2.5
5.0
7.5
Gene count
P value
0.002
0.004
0.006
Cluster 8
Enrichment analysis by Enrichr
cytokine-mediated signaling pathway (GO:0019221)
cellular response to interferon-gamma (GO:0071346)
interferon-gamma-mediated signaling pathway (GO:00...
antigen processing and presentation of exogenous p...
antigen processing and presentation of peptide ant...
antigen processing and presentation of exogenous p...
protein targeting to ER (GO:0045047)
antigen receptor-mediated signaling pathway (GO:00...
SRP-dependent cotranslational protein targeting to...
cytoplasmic translation (GO:0002181)
cotranslational protein targeting to membrane (GO:...
positive regulation of T cell proliferation (GO:00...
cellular response to cytokine stimulus (GO:0071345...
peptide antigen assembly with MHC protein complex ...
nuclear-transcribed mRNA catabolic process, nonsen...
T cell receptor signaling pathway (GO:0050852)
neutrophil activation involved in immune response ...
negative regulation of lymphocyte proliferation (G...
positive regulation of cytokine production (GO:000...
neutrophil chemotaxis (GO:0030593)
Enriched terms
0
5
10
15
20
25
Gene count
P value
5.0e-08
1.0e-07
1.5e-07
2.0e-07
2.5e-07
