## Supplemental Data for "Single-cell RNA sequencing identifies subtypes of cancer-associated fibroblasts in early and late stages of mycosis fungoides"

**Conflict of Interest Statement:** None to disclose

**Running Title:** *ScRNA sequencing identifies CAF subtypes in MF*

**Abbreviations**:
scRNA – single-cell RNA
CAF – Cancer-associated fibroblasts
MF – mycosis fungoides

**Reprint requests:** Courtney Johnson

**Manuscript word count**: 1933 words (2500-word limit)
**Abstract**: 250 words (250-word structured text)
**Translational Relevance:** 112 (120–150-word statement)
**References**: 22 (20 limit)
**Figures and Tables: 3** (4 tables and/or figures limit)

**Keywords:** medical dermatology; cutaneous lymphoma; mycosis fungoides; tumor microenvironment; single-cell sequencing; cancer-associated fibroblasts

**Supplemental Methods and Materials:**

Patient sample. The Johns Hopkins Institutional Review Board approved the study protocol for specimen collection and sequencing (IRB00331637). With informed consent, four MF patients and one without MF were recruited from the Johns Hopkins Dermatology Cutaneous Lymphoma Clinic. All patients diagnosed with MF had biopsy-proven disease, as confirmed by a dermatopathologist. For scRNA-seq, thick plaques or tumors were selected, and a 4 mm punch size was used to collect tissue. For the patient without CTCL, normal, unaffected skin from a double-covered truncal location was selected for collection. Disease staging for patients with MF was determined by EORTC/WHO staging guidelines, in which patients with early-stage MF were defined as stage IA to IIA and late-stage MF as IIB or higher. Freshly dissociated skin samples were transported in cold RPMI + 10% FBS media on ice for immediate tissue dissociation.

Tissue dissociation and single-cell suspension preparation. Freshly collected tissue was washed twice with 10 mL of RPMI + 10% FBS media, and subcutaneous fat was removed. To obtain single-cell suspensions, the samples were transferred to a gentleMACS C tube (Miltenyi Biotec, 130-093-237) containing a solution of 435 µL Buffer L, 12.5 µL Enzyme P, 10 µL Enzyme D, and 12.5 µL Enzyme A from the Whole Skin Dissociation Kit (Miltenyi Biotec, 130-101-540). The sample and enzymes were incubated in a water bath for one hour at 37° C. Following incubation, the sample and enzymes were diluted with 500ml of ice-cold RPMI + 10% FBS media. The C tube was placed onto a gentleMACS dissociator (Miltenyi Biotec, 130-095-937) for 37 seconds on the pre-programmed gentleMACS program h_skin_01 program. After program termination, the C tube was detached, and a short 10-second centrifugation step at the highest speed was performed to collect the sample material at the bottom of the tube. The cell suspension was filtered using a 70 µm pre-wetted filter using 10 mL of cold RPMI + 10% FBS media. The cell suspension was centrifuged at 300 x G for 10 minutes at 4°C. The supernatant was removed entirely. The remaining cells were resuspended in 3100 µL of ice-cold PBS and 900 µL of cold Debris Removal solution (Miltenyi Biotec, 130-109-398). Following slow mixing ten times with a pipette, 4 mL of ice-cold PBS was added to the cells. Then, the suspension was centrifuged at 3000 x G for 10 mins at 4°C. Following centrifugation, the top two debris phases were discarded. The remaining cell suspension was filled to a final volume of 15mL with ice-cold PBS buffer, tightly covered with a cap, and gently inverted three times. Cells were centrifuged at 1000 x G at 4°C for 10 minutes with full acceleration and full brake. The supernatant was aspirated entirely. Cells were resuspended in 500 µL of ice-cold RPMI+10% FBS media for cell viability assessment with trypan blue and cell counting using a hemocytometer. Isolated cells were diluted in ice-cold PBS containing 0.04% BSA to a volume of 700-1200 cells/µL.

Single-cell capture, library preparation, and RNA sequencing. Cell suspensions of 10,000 cells were loaded per lane on a 10X Chromium microfluidic chip and placed into the 10X Chromium controller, according to the manufacturer's protocol (CG000315 Rev C). Subsequent single-cell capture, barcoding, and library preparation were performed using the 10X Chromium Next GEM Single Cell 3' Kit, version 3.1 (Dual index). cDNA and libraries were checked for quality on Agilent Tapestation and quantified on the Qubit 3 Fluorometer. All samples were combined and loaded onto an SP flow cell for sequencing on an Illumina NovaSeq SP sequencer to an average depth of 40-60 million reads per sample.

Pre-processing and quality control of ScRNA-seq data. ScRNA-seq data were demultiplexed and converted to FASTQ format by 'CellRanger mkfastq' (10x Genomics, cellranger-7.0.0), and scRNA-seq files were aligned to the hg19 reference genome using 'CellRanger count' (10x Genomics, cellranger-7.0.0). Using R Studio (R version 4.3.1), cell ranger output data files consisting of barcodes.tsv.gz, features.tsv.gz, and matrix.mtx.gz were loaded into Seurat (4.4.0) to create a sparse count matrix object with dimensions of genes by cells. The *CreateSeruatObject* function was applied to each count matrix to create a Seurat Object for each sample. Quality control (QC) metrics were applied to include only cells with gene counts >200 or <7500 and mitochondrial percentages <5%. The *ScTransform* (SCT) function was applied independently to each post-QC Seurat object, regressing out percent mitochondrial genes and percent ribosomal genes. The SCT performs a variance-stabilizing normalization, accounting for sequencing depth and technical noise (percent.mt and percent.ribo).

Merging and harmonization of data: The *SelectIntegrationFeatures* function was used to identify the top 3000 most highly expressed genes in each Seurat object. All five SCT normalized samples were merged using the *merged_seurat* feature to create one Seurat Object of all patient samples using the harmonized Seurat function. On this newly merged SeuratObject, the most variable gene features were identified, and the top 50 principal components were calculated using the *RunPCA* function. The *RunHarmony* integration function was applied to the merged SeuratObject to determine each cluster's maximum diversity and batch correct cells based on its principal component coordinates [11, 12]. A new reduction of 50 “harmonized components” was added to the merged Seurat object, and a UMAP of 1:30 dimensions was created from the 50 harmonized PCs. *FindNeighbors* (reduction set to harmony) and *FindClusters* (resolution of 0.2, 0.4, 0.6, 0.8, 1.0, and 1.2) functions were performed to determine clusters. Cell clusters were visualized using UMAP (Seurat package 4.4.0, R version 4.3.1).

Cell type annotation and statistical analysis: Differential gene expression was conducted on each cluster using the *FindMarkers* function (Wilcoxon Rank Sum Test) on the RNA assay within the Seurat Object. Genes were considered significant markers if they were expressed in at least 10–25% of cells in the tested groups (min.pct filter) and exhibited an absolute log fold-change greater than 0.25 (logfc.threshold). Statistical significance was determined using the Wilcoxon rank-sum test, and resulting *p*-values were adjusted for multiple testing using the Benjamini–Hochberg procedure. Only genes with an adjusted *p*-value <0.05 meeting these criteria were retained as cluster-specific markers. Cluster-specific markers identified as up-regulated in multiple clusters were disregarded, and only cluster-specific differentially expressed genes were retained. Canonical gene markers were used to annotate the clusters and to name each cluster type (**Supplemental Table S2**). Fibroblast cells, identified by expression of *COL1A1,* were subset for further analysis and clustered based on the top 3000 variable genes **(Supplemental Figure S2).**

**Supplemental Legends:**

**Supplemental Table S1. Sequencing and quality control metrics.** Quality control thresholds included removal of cells with < 200 genes or > 15% mitochondrial transcripts. Total cell counts and median gene expression per sample.

**Supplemental Table S2. Canonical gene markers are used to identify major cell populations.** List of canonical marker genes applied for annotation of transcriptional clusters in the combined dataset of mycosis fungoides (MF) and healthy control (HC) skin. Markers include fibroblasts (COL1A1), T cells (CD3D), natural killer T cells (NKG7), keratinocytes (KRT1, KRT14, KRT5), monocytes/macrophages (AIF1, FCER1G), endothelial cells (PECAM1, VWF), B cells (MS4A1), plasma cells (JCHAIN), mitotic T cells (CD3D, MK167), mast cells (TPSB2, FCER1G), conventional dendritic cells (CD14), and plasmacytoid dendritic cells (IL3RA). Clusters were annotated based on the highest average expression of these markers following log-normalization and scaling in Seurat v4.

**Supplemental Figure S1. Unsupervised clustering of all cell types from mycosis fungoides (MF) and healthy control (HC) skin.** Uniform Manifold Approximation and Projection (UMAP) visualization reveals 29 transcriptionally distinct clusters derived from the combined dataset of HC and MF skin cells. Clustering was performed using the Seurat v4 pipeline, which incorporates principal component analysis-based dimensionality reduction and shared nearest neighbor modularity optimization (resolution = 0.5). Cluster identities were assigned using canonical marker genes for keratinocytes (KRT1, KRT14, KRT5), fibroblasts (COL1A1), T cells (CD3D), endothelial cells (PECAM1, VWF), B cells (MS4A1), plasma cells (JCHAIN), natural killer T cells (NKG7), monocytes/macrophages (AIF1, FCER1G), mast cells (TPSB2, FCER1G), and dendritic cell subsets (CD14, IL3RA). Differential expression was calculated using the Wilcoxon rank-sum test with Bonferroni correction (adjusted $p < 0.05$).

**Supplemental Figure S2. Heatmap of the top five differentially expressed genes per cluster in the combined dataset.**
Heatmap showing the top five differentially expressed genes (DEGs) for each transcriptional cluster across all cells from mycosis fungoides (MF) and healthy control (HC) skin. Expression values are scaled per gene (z-score). Clustering was performed on log-normalized counts using principal component analysis followed by shared nearest neighbor modularity optimization (resolution = 0.5). DEGs were identified using the Wilcoxon rank-sum test implemented in Seurat ($\log_2$ fold-change ≥ 0.25, adjusted $p < 0.05$, Bonferroni correction).

**Supplemental Figure S3. Unsupervised clustering of all cell types separated by patient sample.** Clustering was performed in Seurat v4 using principal component analysis-based dimensionality reduction and shared nearest neighbor modularity optimization (resolution = 0.5). Differential expression was calculated using the Wilcoxon rank-sum test with Bonferroni correction (adjusted $p < 0.05$).

A) Uniform manifold approximation and projection (UMAP) visualization of all identified clusters split by individual patient samples (n = 5; 1 healthy control and 4 mycosis fungoides (MF)). Cell identities were assigned using canonical markers for fibroblasts (COL1A1), T cells (CD3D), keratinocytes (KRT1, KRT14, KRT5), natural killer T cells (NKG7), monocytes/macrophages (AIF1, FCER1G), endothelial cells (PECAM1, VWF), B cells (MS4A1), plasma cells (JCHAIN), mitotic T cells (CD3D, MK167), mast cells (TPSB2, FCER1G), and dendritic cell subsets (CD14, IL3RA).
B) Combined UMAP projection colored by sample ID showing integration across patients.

**Supplemental Figure S4. Heatmap of the top five differentially expressed genes within each fibroblast subcluster.**
Heatmap showing the top five differentially expressed genes (DEGs) for each fibroblast subcluster derived from mycosis fungoides (MF) and healthy control (HC) skin samples. Expression values are log-normalized counts scaled per gene (z-score). The distinct expression signatures confirm the presence of unique fibroblast subpopulations. DEGs were identified using the Wilcoxon rank-sum test ($log_2$ fold-change ≥ 0.25, adjusted $p < 0.05$, Bonferroni correction).

**Supplemental Figure S5. UMAP visualization of fibroblast subclusters by patient sample.** Clustering was performed on COL1A1$^+$ cells using principal component analysis and shared nearest neighbor modularity optimization (resolution = 0.6). Differential expression analysis was conducted with the Wilcoxon rank-sum test ($log_2$ fold-change ≥ 0.25, adjusted $p < 0.05$, Bonferroni correction).
A) Uniform manifold approximation and projection (UMAP) of COL1A1$^+$ fibroblast subclusters split by individual patient samples (n = 5; 1 healthy control and 4 mycosis fungoides [MF]). Nine transcriptionally distinct fibroblast subclusters are color-coded.
B) Combined UMAP projection of all fibroblast cells colored by sample ID showing integration across patients.

**Supplemental Figure S6. Gene ontology (GO) enrichment analysis of fibroblast subclusters.**
Bar plots showing the top enriched GO biological process terms for differentially expressed genes (DEGs) in each fibroblast subcluster (clusters 0–8). GO analysis highlights functional heterogeneity among fibroblast subpopulations, including extracellular matrix organization, cytokine signaling, mitochondrial metabolism, immune modulation, and vascular development pathways. DEGs were identified using the Wilcoxon rank-sum test ($log_2$ fold-change ≥ 0.25, adjusted $p < 0.05$, Bonferroni correction). GO enrichment analysis was performed using the top 50 DEGs per cluster via a hypergeometric test with Benjamini–Hochberg correction ($q < 0.05$).

**Additional References:**


21. Querfeld C, Leung S, Myskowski PL, Curran SA, Goldman DA, Heller G, Wu X, Kil SH, Sharma S, Finn KJ, Horwitz S, Moskowitz A, Mehrara B, Rosen ST, Halpern AC, Young JW: **Primary T Cells from Cutaneous T-cell Lymphoma Skin Explants Display an Exhausted Immune Checkpoint Profile.** Cancer immunology research 2018, **6**(8):900–909.
22. Chen X, Chen F, Jia S, Lu Q, Zhao M: **Antigen-presenting fibroblasts: emerging players in immune modulation and therapeutic targets.** Theranostics 2025, **15**(8):3332–3344.
23. O'Connor RA, Martinez BR, Koppensteiner L, Mathieson L, Akram AR: **Cancer-associated fibroblasts drive CXCL13 production in activated T cells via TGF-beta.** Frontiers in immunology 2023, **14**:1221532.
24. Yan X, Orentas RJ, Johnson BD: **Tumor-derived macrophage migration inhibitory factor (MIF) inhibits T lymphocyte activation.** Cytokine (Philadelphia, Pa.) 2006, **33**(4):188–198.
25. Lin C, Chen Y, Zhang F, Zhu P, Yu L, Chen W: **Single-cell RNA sequencing reveals the mediatory role of cancer-associated fibroblast PTN in hepatitis B virus cirrhosis-HCC progression.** Gut Pathog 2023, **15**(1):26–z.
26. Shao M, Wang L, Zhang Q, Wang T, Wang S: **STMN2 overexpression promotes cell proliferation and EMT in pancreatic cancer mediated by WNT/beta-catenin signaling.** Cancer Gene Ther 2023, **30**(3):472–480.
27. Lee HJ, Kwak Y, Na YS, Kim H, Park MR, Jo JY, Kim JY, Cho S, Kim P: **Proteomic Heterogeneity of the Extracellular Matrix Identifies Histologic Subtype-Specific Fibroblast in Gastric Cancer.** Mol Cell Proteomics 2024, **23**(10):100843.
28. Chu P, Tzeng YT, Tsui K, Chu C, Li C: **Downregulation of ATP binding cassette subfamily a member 10 acts as a prognostic factor associated with immune infiltration in breast cancer.** Aging (Albany NY) 2022, **14**(5):2252–2267.
29. Hao Q, Zhou X: **The emerging role of long noncoding RNA RMRP in cancer development and targeted therapy.** Cancer Biol Med 2022, **19**(2):140–146.
30. Picchio MC, Scala E, Pomponi D, Caprini E, Frontani M, Angelucci I, Mangoni A, Lazzeri C, Perez M, Remotti D, Bonoldi E, Benucci R, Baliva G, Lombardo GA, Napolitano M, Russo G, Narducci MG: **CXCL13 is highly produced by Sezary cells and enhances their migratory ability via a synergistic mechanism involving CCL19 and CCL21 chemokines.** Cancer Res 2008, **68**(17):7137–7146.
31. Lounnas N, Rosilio C, Nebout M, Mary D, Griessinger E, Neffati Z, Chiche J, Spits H, Hagenbeek TJ, Asnafi V, Poulsen S, Supuran CT, Peyron J, Imbert V: **Pharmacological inhibition of carbonic anhydrase XII interferes with cell proliferation and induces cell apoptosis in T-cell lymphomas.** Cancer Lett 2013, **333**(1):76–88.
32. Wang H, Chen J, Chen X, Liu Y, Wang J, Meng Q, Wang H, He Y, Song Y, Li J, Ju Z, Xiao P, Qian J, Song Z: **Cancer-Associated Fibroblasts Expressing Sulfatase 1 Facilitate VEGFA-Dependent Microenvironmental Remodeling to Support Colorectal Cancer.** Cancer Res 2024, **84**(20):3371–3387.
33. El-Ashmawy AA, Shamloula MM, Elfar NN: **Podoplanin as a Predictive Marker for Identification of High-Risk Mycosis Fungoides Patients: An Immunohistochemical Study.** Indian J Dermatol 2020, **65**(6):500–505.

34. Zhan S, Li J, Ge W: **Multifaceted Roles of Asporin in Cancer: Current Understanding.** Front Oncol 2019, **9**:948.
35. Wu Y, Huang Y, Chang T, Chen C, Wu P, Huang S, Chou C: **COL11A1 activates cancer-associated fibroblasts by modulating TGF-beta3 through the NF-kappaB/IGFBP2 axis in ovarian cancer cells.** Oncogene 2021, **40**(26):4503–4519.
36. Zhou X, Han J, Zuo A, Ba Y, Liu S, Xu H, Zhang Y, Weng S, Zhou Z, Liu L, Luo P, Cheng Q, Zhang C, Chen Y, Shan D, Liu B, Yang S, Han X, Deng J, Liu Z: **THBS2 + cancer-associated fibroblasts promote EMT leading to oxaliplatin resistance via COL8A1-mediated PI3K/AKT activation in colorectal cancer.** Mol Cancer 2024, **23**(1):282–y.
37. Liu X, Xing C: **PDZRN4-mediated colon cancer cell proliferation and dissemination is regulated by miR-221-3p.** Transl Cancer Res 2019, **8**(4):1289–1300.
38. Huang Y, Zhang Y, Shen Z, Xu J, Sheng J: **FHOD3 shows clinical significance in progression of ovarian cancer through regulation of caspase-3 signaling pathway.** Gene 2025, **933**:148943.
39. Jiang M, Zhao F, Lou T: **Assessment of Significant Pathway Signaling and Prognostic Value of GNG11 in Ovarian Serous Cystadenocarcinoma.** Int J Gen Med 2021, **14**:2329–2341.
40. Tarnowski M, Grymula K, Liu R, Tarnowska J, Drukala J, Ratajczak J, Mitchell RA, Ratajczak MZ, Kucia M: **Macrophage migration inhibitory factor is secreted by rhabdomyosarcoma cells, modulates tumor metastasis by binding to CXCR4 and CXCR7 receptors and inhibits recruitment of cancer-associated fibroblasts.** Mol Cancer Res 2010, **8**(10):1328–1343.
41. Wang J, Akter R, Shahriar MF, Uddin MN: **Cancer-Associated Stromal Fibroblast-Derived Transcriptomes Predict Poor Clinical Outcomes and Immunosuppression in Colon Cancer.** Pathol Oncol Res 2022, **28**:1610350.
42. Kim H, Chung H, Yoo Y, Kim H, Lee J, Lee M, Kong G: **Inhibitor of DNA binding 1 activates vascular endothelial growth factor through enhancing the stability and activity of hypoxia-inducible factor-1alpha.** Mol Cancer Res 2007, **5**(4):321–329.
43. Zhu H, Zhang X, Gu C, Zhong Y, Long T, Ma Y, Hu Z, Li Z, Wang X: **Cancer-associated fibroblasts promote colorectal cancer progression by secreting CLEC3B.** Cancer Biol Ther 2019, **20**(7):967–978.
44. Guo Q, Gao X, Li J, Liu Y, Liu J, Yang H, Cui M, Zhang M, Duan L, Ma X: **High expression of PCOLCE gene indicate poor prognosis in patients and are associated with immune infiltration in glioma.** Sci Rep 2023, **13**(1):3820–5.
45. Yu Y, Wang Z, Zheng Q, Li J: **GREB1L overexpression correlates with prognosis and immune cell infiltration in lung adenocarcinoma.** Sci Rep 2021, **11**:13281.
46. Dasgupta S, Ghosh T, Dhar J, Bhuniya A, Nandi P, Das A, Saha A, Das J, Guha I, Banerjee S, Chakravarti M, Dasgupta PS, Alam N, Chakrabarti J, Majumdar S, Chakrabarti P, Storkus WJ, Baral R, Bose A: **RGS5-TGFbeta-Smad2/3 axis switches pro- to anti-apoptotic signaling in tumor-residing pericytes, assisting tumor growth.** Cell Death Differ 2021, **28**(11):3052–3076.
47. Kayamori K, Katsube K, Sakamoto K, Ohyama Y, Hirai H, Yukimori A, Ohata Y, Akashi T, Saitoh M, Harada K, Harada H, Yamaguchi A: **NOTCH3 Is Induced in**

**Cancer-Associated Fibroblasts and Promotes Angiogenesis in Oral Squamous Cell Carcinoma.** PLoS One 2016, **11**(4):e0154112.
48. Chen Z, Wei X, Wang X, Zheng X, Chang B, Shen L, Zhu H, Yang M, Li S, Zheng X: **NDUFA4L2 promotes glioblastoma progression, is associated with poor survival, and can be effectively targeted by apatinib.** Cell Death Dis 2021, **12**(4):377–3.
49. Vasiukov G, Zou Y, Senosain M, Rahman JSM, Antic S, Young KM, Grogan EL, Kammer MN, Maldonado F, Reinhart-King CA, Massion PP: **Cancer-associated fibroblasts in early-stage lung adenocarcinoma correlate with tumor aggressiveness.** Sci Rep 2023, **13**(1):17604–3.
50. Mauri F, Schepkens C, Lapouge G, Drogat B, Song Y, Pastushenko I, Rorive S, Blondeau J, Golstein S, Bareche Y, Miglianico M, Nkusi E, Rozzi M, Moers V, Brisebarre A, Raphaël M, Dubois C, Allard J, Durdu B, Ribeiro F, Sotiriou C, Salmon I, Vakili J, Blanpain C: **NR2F2 controls malignant squamous cell carcinoma state by promoting stemness and invasion and repressing differentiation.** Nat Cancer 2021, **2**(11):1152–1169.
51. Zhu L, Yu Q, Li Y, Zhang M, Peng Z, Wang S, Quan Z, Gao D: **SKAP1 Is a Novel Biomarker and Therapeutic Target for Gastric Cancer: Evidence from Expression, Functional, and Bioinformatic Analyses.** Int J Mol Sci 2023, **24**(14):11870. doi: 10.3390/ijms241411870.
52. Geng Y, Liu X, Liang J, Habiel DM, Kulur V, Coelho AL, Deng N, Xie T, Wang Y, Liu N, Huang G, Kurkciyan A, Liu Z, Tang J, Hogaboam CM, Jiang D, Noble PW: **PD-L1 on invasive fibroblasts drives fibrosis in a humanized model of idiopathic pulmonary fibrosis.** JCI Insight 2019, **4**(6):e125326. doi: 10.1172/jci.insight.125326. eCollection 2019 Mar 21.
53. Cressey R, Han MTT, Khaodee W, Xiyuan G, Qing Y: **Navigating PRKCSH's impact on cancer: from N-linked glycosylation to death pathway and anti-tumor immunity.** Front Oncol 2024, **14**:1378694.
54. Xu C, Fang H, Gu Y, Yu K, Wang J, Lin C, Zhang H, Li H, He H, Liu H, Li R: **Impact of intratumoural CD96 expression on clinical outcome and therapeutic benefit in gastric cancer.** Cancer Sci 2022, **113**(12):4070–4081.
55. Fils-Aime N, Dai M, Guo J, El-Mousawi M, Kahramangil B, Neel J, Lebrun J: **MicroRNA-584 and the protein phosphatase and actin regulator 1 (PHACTR1), a new signaling route through which transforming growth factor-beta Mediates the migration and actin dynamics of breast cancer cells.** J Biol Chem 2013, **288**(17):11807–11823.
56. Velmurugan BK, Yeh K, Hsieh M, Yeh C, Lin C, Kao C, Huang L, Lin S: **UNC13C Suppress Tumor Progression via Inhibiting EMT Pathway and Improves Survival in Oral Squamous Cell Carcinoma.** Front Oncol 2019, **9**:728.
57. Sun Y, Preiss NK, Valenteros KB, Kamal Y, Usherwood Y, Frost HR, Usherwood EJ: **Zbtb20 Restrains CD8 T Cell Immunometabolism and Restricts Memory Differentiation and Antitumor Immunity.** J Immunol 2020, **205**(10):2649–2666.
58. Brazzelli V, Rivetti N, Badulli C, Carugno A, Grasso V, De Silvestri A, Martinetti M, Borroni G: **Immunogenetic factors in mycosis fungoides: can the HLA system influence the susceptibility and prognosis of the disease? Long-term follow-up study of 46 patients.** J Eur Acad Dermatol Venereol 2014, **28**(12):1732–1737.

59. Shi K, Li Q, Zhang Y, Huang H, Ding D, Luo W, Zhang J, Guo Q: **HLA-DPA1 overexpression inhibits cancer progression, reduces resistance to cisplatin, and correlates with increased immune infiltration in lung adenocarcinoma.** Aging (Albany NY) 2023, **15**(20):11067–11091.
60. Zhou J, Xie T, Shan H, Cheng G: **HLA-DQA1 expression is associated with prognosis and predictable with radiomics in breast cancer.** Radiat Oncol 2023, **18**(1):117–4.
